# Brain derived neurotrophic factor (BDNF) Val66Met polymorphism is not risk factor for bipolar disorder

**DOI:** 10.1101/19010280

**Authors:** Vandana Rai, Farhin Jamal, Pradeep Kumar

## Abstract

Bipolar disorder (BPD) is a psychiatric disease, characterized by the cycles of mania and depression. Several genetic studies investigated BDNF gene Val66Met polymorphism as risk factor for BPD, but results were inconclusive. Therefore, present meta-analysis was performed to reevaluate the BDNF Val66Met polymorphism and BPD association. Four databases (Pubmed, Springer Link, Science Direct and Google Scholar) were searched for eligible studies up to March 31,2018. Pooled odds ratios (OR) with 95% confidence intervals (CI) were calculated to estimate the strength of the association. All statistical analyses were done by MetaAnalyst and Mix program. Forty studies with a total of 28,787 subjects (10,085 cases and 18,702 controls) were included in this meta-analysis. Overall, pooled analysis indicated that there was no significant association between BDNF Val66Met polymorphism and BPD risk under all five genetic models (OR_A vs.G_ =0.99, 95%CI= 0.94-1.03, p=0.49; OR_AG vs. GG_= 0.1.02, 95%CI= 0.95-1.07, p= 0.57; OR_AA vs. GG_ = 0.98, 95%CI=0.89-1.08, p=0.75; OR_AA+AG vs. GG_= 1.0, 95%CI= 0.94-1.06, p= 0.89;OR_AA vs. AG+GG_= 0.96, 95%CI= 0.89-1.05, p= 0.47). Similarly, no significant association was observed in ethnicity based subgroup analysis in both Asian and Caucasian population. However, significant association was found in subtype analysis between BDNF Val66Met and BPDII (OR_AA+AG vs. GG_= 1.21, 95%CI= 1.06-1.37, p= 0.003) but not with BPDI. These findings suggested that the BDNF Val66Met polymorphism confer no genetic susceptibility to BPD I but risk for BPDII.

## Introduction

Bipolar disorder (BPD) is one of the most common and severe psychiatric disorder, which is classified as a mood disorder in the Diagnostic and statistical Manual of Mental Disorders(DSM IV) (American Psychiatric Association,2000). World Health Organization has identified BPD as the sixth leading cause of disability in the world(WHO,2004). The lifetime prevalence of BPD is estimated to be between 1% and 2% (Kessler et al.,1994; Merikangas et al.,2007). The disorder is characterized by episodes of mania, with elated or irritable-angry mood and symptoms like pressured speech, racing thoughts, grandiose ideas, increased energy, and reckless behavior, alternating with more normal periods and, in most cases, with episodes of depression. Although the etiologic mechanisms for BPD are not well known, it has been hypothesized that both environmental and genetic factors play important roles, and multiple genes contribute to risk of the disease (Hawton et al.,2005; Goodwin,20012).

BDNF is one of the most studied and abundant neurotrophin in central nervous system, which plays an important role in a variety of neural processes. During, BDNF is essential for neurogenesis, neuronal survival, and neural development pathways. In the adult, BDNF is important for synaptic plasticity, dendritic growth and long-term memory consolidation (Post,2007). Because of the diverse functions of BDNF protein, BDNF gene is a good candidate for the susceptibility to different neuropsychiatric disorders including BPD (Goldberg et al.,2008).

Several polymorphisms are reported in BDNF gene, but in exon 5 G to A (G196A) transition (dbSNP: rs6265) at position 196 is most studied and clinically important. G196A polymorphism produces amino acid substitution of valine (Val) to methionine (Met) at codon 66(Val66Met) in the 5′ prodomain in the BDNF protein (Cargill et al.,1999). This substitution from valine (Val) to methionine (Met) at codon 66 has functional consequences (Egan etal.,2003; Chen et al.,2006). The Met (A) allele of the precursor peptide has been associated with impaired intracellular trafficking of pro-BDNF into dendrites and vesicles as well as a reduction inactivity-dependent secretion, the process that plays a major role in the regulation of extracellular levels of BDNF (Egan etal.,2003; Chen et al.,2006). The frequencies of the Val and Met alleles of BDNF Val66Met vary by ethnicity; about 80% of the European population has Val allele while only 50% of Asian has it (Pivac et al.,2009). Numerous studies investigated Val66Met polymorphism as a risk factor for BPD but results were inconclusive. Hence, authors performed present meta-analysis to reevaluate the association between BDNF Val66Met polymorphism and BPD risk.

## Methods

Meta-analysis was carried out according to Meta-analysis of observational studies in epidemiology (MOOSE) guidelines (Stroup et al.,2000).

### Search Strategy

A literature search was performed for available articles that were published up to March 31, 2018 from the following databases: (1) Pubmed; (2) Google Scholar; (3)Science Direct; and (4) Springer Link. The search used the following keywords: ‘‘Bipolar Disorder’’ or ‘‘BPD’’ and ‘‘brain-derived neurotrophic factor’’ or ‘‘BDNF’’ and ‘‘Val66Met’’. We also reviewed the bibliography of included articles to identify additional articles not retrieved by database search.

### Inclusion and exclusion criteria

The following inclusion criteria were used: (i) studies should be original, (ii) used case control approach, and (iii) used standard diagnostic criteria for bipolar disorder patient diagnosis (Diagnostic and Statistical Manual of Mental Disorders (DSM-IV) or the International Classification of Diseases (ICD). Studies were excluded if: (i) incomplete raw data information like-number of genotype/allele, and (ii) studies based on pedigree data.

### Data Extraction

Relevant information were extracted from all selected studies like-first author’s name, year of publication, country name, ethnicity, sample size and number of cases and controls for BDNF Val66Met genotypes (GG, AG and AA). Mutant allele frequencies for the cases and controls were calculated from corresponding genotypes. Allele frequency was calculated by simple gene count method.

### Statistical analysis

The present meta-analysis examined the overall association of Met allele with the risk of bipolar disorder. Pooled odds ratio (OR) with 95% confidence interval (CI) was used as association measure, which was estimated either by fixed effect (Mantel and Haenszel,1959) or random effect (DerSimonian and Laird,1968) models depending upon heterogeneity. Heterogeneity was tested by Q-statistics and quantified by the I^2^ statistic (Higgins and Thompson,2002). If I^2^ > 50% then random effect model was used (Whitehead,2002). Genetic models were chosen according to the method of Thakkinstian et al. (2005),, briefly calculating and comparing the ORs of A(Met) vs. G (Val) (allele contrast), AA(Met/Met) vs. GG(Val/Val) (homozygote), AG(Met/Val) vs. GG(Val/Val) (co-dominant), AA(Met/Met) + AG(Met/Val) vs. GG (Val/Val) (dominant) and AA(Met/Met) vs. AG(Met/Val) + GG(Val/Val) (recessive) models. We have also done sub-group analysis based on ethnicity and subtypes of bipolar disorder like BPDI and BPDII. In allele contrast meta-analysis, sensitivity analysis performed by exclusion of the studies in which control population was not in Hardy Weinberg equilibrium and studies with small sample size.

### Publication bias

Publication bias was investigated by funnel plots; viz. funnel plot of standard error by log odds ratio and funnel plot of precision (1/standard error) by log odds ratio. Different statistical tests such as Begg and Mazumdar rank correlation (Begg and Mazumdar,1994) and Egger’s regression intercept (Egger et al.,1997) were adopted to assess the publication bias. All p values were two tailed with a significance level at 0.05. All statistical analyses were undertaken using program MetaAnalyst (Wallace et al.,2013) and MIX version 1.7 (Bax eta al.,2006).

## Results

### Characteristics of included studies

The study search workflow was shown in Figure 1. The preliminary search resulted in 187 publications from Pubmed, Springer Link, Science Direct and Google Scholar. Out of which 104 articles were excluded after title and abstract reading. Again 23 articles including review, letter to editors and comment were excluded from remaining 83 articles. Further 26 articles, which were duplicate studied, studies without genotype or allele number, studies on animal models, were excluded. Out of 34 remaining articles, 3 articles were meta-analysis, and excluded.. Thus, total thirty studies that investigated the association of BDNF Val66Met polymorphism with BPD were found suitable for the inclusion in the present meta-analysis (Hong et al.,2003; Nakata et al., 2003; Kunugi et al., 2004; Oswald et al.,2004; Skibinska et al.,2004; Lohoff et al., 2004; Nerves-Pereira et al., 2005,2011; Schumacher et al., 2005; Green et al., 2006; Liu et al., 2007; Tramontina et al., 2007; Kim et al., 2008; Vincze et al., 2008; Tang et al., 2008; Xu et al., 2009; Hosang et al., 2010; Carrard et al.,2011; Huang et al., 2012; Pae et al., 2012; Wang et al., 2012; Lee et al., 2012; Min et al., 2012; Chang et al., 2013; Lee et al.,2013; Gonzalez-Castro et al., 2014; Frazier et al., 2014; Kenna et al., 2014; Nassan et al., 2015; Morales-Marín et al., 2016). In nine articles, authors (Nakata et al.,2003; Kunugi et al., 2004; Green et al., 2006; Xu et al., 2009; Huang et al., 2012; Wang et al., 2012; Lee et al., 2012; Chang et al., 2013; Kenna et al., 2014) investigated subtypes of bipolar disorder like BPDI and BPD II. We included each case sample group (BPD I and BPD II case samples) as separate article so total forty article were included in the present meta-analysis (Table 1).

**Table 1.** Details of included studies.

| Study | Country | Type of Bipolar disorder | Case no. | Control no. | Case Genotypes |  |  | Control Genotypes |  |  | P- value of HWE |
| --- | --- | --- | --- | --- | --- | --- | --- | --- | --- | --- | --- |
|  |  |  |  |  | Val/Val | Val/Me t | Met/Met | Val/Val | Val/Me t | Met/Me t |  |
| Hong et al 2003 | China | BPD | 192 | 392 | 97 | 212 | 83 | 97 | 212 | 83 | 0.1 |
| Nakata et al.,2003 | Japan | BPDI | 100 | 190 | 63 | 94 | 33 | 63 | 94 | 33 | 0.83 |
| Nakata et al.,2003 | Japan | BPDII | 30 | 190 | 63 | 94 | 33 | 63 | 94 | 33 | 0.83 |
| Kunugi et al.,2004 | Japan | BPDI | 347 | 588 | 216 | 270 | 102 | 216 | 270 | 102 | 0.26 |
| Kunugi et al.,2004 | Japan | BPDII | 172 | 588 | 216 | 270 | 102 | 216 | 270 | 102 | 0.26 |
| Oswald et al.,2004 | Bulgaria | BPD | 108 | 158 | 96 | 57 | 5 | 96 | 57 | 5 | 0.31 |
| Skibinska et al.,2004 | Wielkopolaska | BPD | 352 | 375 | 248 | 117 | 10 | 248 | 117 | 10 | 0.38 |
| Lohoff et al.,2005,BPI | USA | BPI | 621 | 1001 | 619 | 338 | 44 | 619 | 338 | 44 | 0.8 |
| Nerves- Pereira et al.,2005 | Scotland | BPD | 263 | 350 | 208 | 131 | 11 | 208 | 131 | 11 | 0.07 |
| Schumacher et al.,2005 | Germany | BPD | 281 | 1097 | 713 | 351 | 33 | 713 | 351 | 33 | 0.19 |
| Green et al.,2006 | UK | BPDI | 864 | 2100 | 1372 | 660 | 68 | 1372 | 660 | 68 | 0.29 |
| Green et al.,2006 | UK | BPDII | 98 | 2100 | 1372 | 660 | 68 | 1372 | 660 | 68 | 0.29 |
| Green et al.,2006 | UK | Rapid Cycle BPD | 131 | 2100 | 1372 | 660 | 68 | 1372 | 660 | 68 | 0.29 |
| Liu et al.,2007 | China | BPD | 100 | 100 | 24 | 51 | 25 | 24 | 51 | 25 | 0.84 |
| Tramontina et al.,2007 | Brazil | BPDI | 114 | 137 | 95 | 40 | 2 | 95 | 40 | 2 | 0.33 |
| Kim et al.,2008 | Korea | BPD | 169 | 251 | 77 | 114 | 60 | 77 | 114 | 60 | 0.16 |
| Vincze et al.,2008 | France | BPD | 336 | 313 | 202 | 128 | 6 | 181 | 111 | 21 | 0.48 |
| Tang et al.,2008 | China | BPD | 197 | 208 | 73 | 92 | 32 | 65 | 105 | 38 | 0.69 |
| Xu et al., 2009 | China | BPDI | 416 | 501 | 135 | 181 | 100 | 141 | 264 | 96 | 0.16 |
| Xu et al.,2009 | China | BPII | 82 | 501 | 19 | 36 | 27 | 141 | 264 | 96 | 0.16 |
| Hosang et al.,2010 | UK | BPD | 488 | 598 | 298 | 184 | 6 | 407 | 169 | 22 | 0.3 |
| Neves et al.,2011 | Brazil | BPD | 152 | 160 | 119 | 31 | 2 | 134 | 21 | 5 | 0.001 |
| Carrad et al.,2011 | Switzerland | BPD | 109 | 158 | 64 | 42 | 3 | 93 | 56 | 9 | 0.88 |
| Huang et al.,2012 | Taiwan | BPDI | 208 | 255 | 44 | 100 | 64 | 61 | 127 | 67 | 0.97 |
| Huang et al.,2012 | Taiwan | BPDII | 329 | 255 | 81 | 177 | 71 | 61 | 127 | 67 | 0.97 |
| Pae et al.,2012 | Korea | BPD | 132 | 170 | 40 | 70 | 22 | 58 | 81 | 31 | 0.77 |
| Wang et al.,2012,BPI | China | BPI | 281 | 386 | 71 | 146 | 64 | 126 | 184 | 76 | 0.55 |
| Wang et al., 2012,BPII | China | BPII | 56 | 576 | 11 | 31 | 14 | 184 | 76 | 316 | 0 |
| Lee et al.,2012 | China | BPI | 268 | 260 | 64 | 130 | 74 | 65 | 127 | 68 | 0.71 |
| Lee et al.,2012 | China | BPII | 436 | 260 | 102 | 230 | 104 | 65 | 127 | 68 | 0.71 |
| Min et al.,2012 | Korea | BPD | 184 | 214 | 66 | 90 | 28 | 73 | 99 | 42 | 0.42 |
| Chang et al., 2013,BPI | China | BPI | 286 | 349 | 81 | 132 | 73 | 99 | 163 | 87 | 0.22 |
| Chang et al.,2013,BPII | China | BPII | 681 | 349 | 156 | 356 | 169 | 99 | 163 | 87 | 0.22 |
| Lee et al.,2013 | China | BPII | 431 | 340 | 221 | 178 | 32 | 194 | 116 | 30 | 0.04 |
| Castro TB et al.,2014 | Mexico | BPI | 139 | 141 | 97 | 38 | 4 | 94 | 42 | 5 | 0.9 |
| Frazier et al.,2014 | USA | BPD | 102 | 135 | 75 | 22 | 5 | 85 | 41 | 9 | 0.19 |
| Kenna et al.,2014 | USA | BPI | 12 | 16 | 9 | 3 | 0 | 10 | 5 | 1 | 0.73 |
| Kenna et al.,2014 | USA | BPII | 14 | 16 | 7 | 7 | 0 | 10 | 5 | 1 | 0.73 |
| Naasan et al.,2015 | USA | BPD | 725 | 764 | 459 | 232 | 34 | 513 | 228 | 23 | 0.7 |
| Morales-Marín et al.,2016 | Mexico | BPD | 79 | 60 | 63 | 16 | 0 | 34 | 23 | 3 | 0.72 |

**Figure 1.**
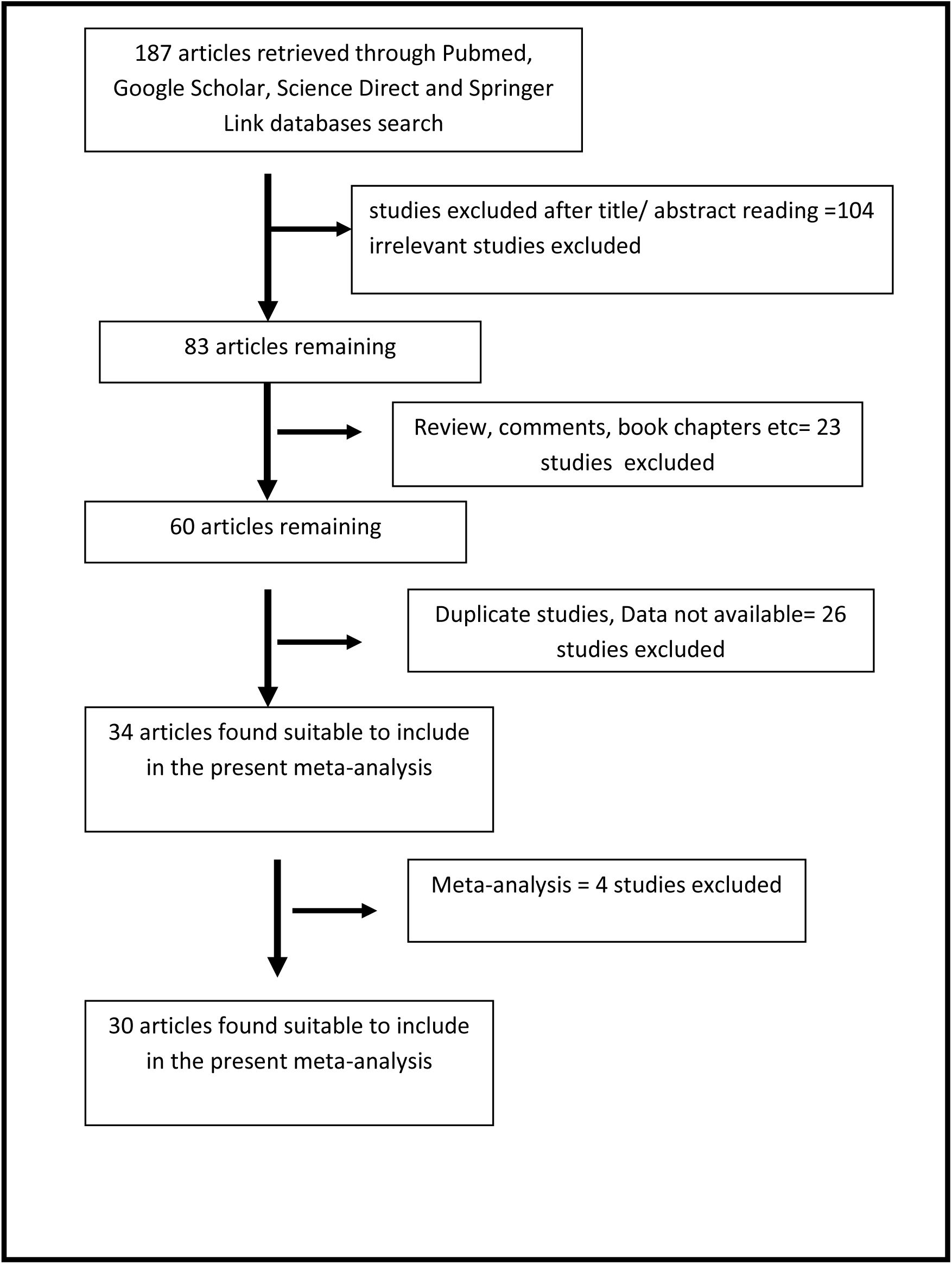
Flow Diagram of Study Search and Selection Process.

In forty included studies, the smallest case sample size was 26 (Kenne et al., 2014) and highest sample size was 864 (Green et al., 2006). In included studies, total cases and controls were 10,085 and 18,702 respectively. In controls genotype percentage of GG, AG and AA were 47.73%, 40.09% and 12.18% respectively. In cases genotype percentage of GG, AG and AA were 52.63%, 36.59% and 10.23% respectively. Control population of three studies (Nerves-Pereira et al.,2011; Wang et al.,2012; Lee et al.,2013) was not in Hardy-Weinberg equilibrium (Table 1). Case and control subjects were selected from following countries-Brazil (Tramontina et al., 2007; Nerves et al., 2011), Bulgaria (Oswald et al.,2004), China (Hong et al.,2003; Liu et al., 2007; Tang et al., 2008; Xu et al., 2009; Wang et al., 2012; Lee et al., 2012; Chang et al., 2013; Lee et al.,2013), France (Vincze et al., 2008), Germany (Schumacher et al., 2005), Japan (Nakata et al., 2003; Kunugi et al., 2004), Korea (Kim et al., 2008; Pae et al., 2012; Min et al., 2012), Mexico (Gonzalez-Castro et al., 2014; Morales-Marín et al., 2016), Scotland (Nerves-Pereira et al., 2005), Taiwan (Huang et al., 2012), UK (Green et al., 2006; Hosang et al., 2010), USA (Lohoff et al., 2004; Frazier et al., 2014; Kenna et al., 2014; Nassan et al., 2015),Wielkopolaska (Skibinska et al.,2004).

### Meta-analysis

Meta-analysis with allele contrast (A (Met) vs. G(Val)) showed no association with both fixed effect (OR_A vs.G_=0.99, 95%CI= 0.94-1.03, p=0.49) and random effect model (OR_A vs.G_ =0.98, 95%CI= 0.93-1.03, p=0.46)(Table 2; Figure 2). Significant association was not found between BPD and mutant genotype (AA vs. GG; homozygote model) with both fixed (OR_AA vs. GG_ = 0.98, 95%CI=0.89-1.08, p=0.75) and random (OR_AA vs. GG_ =0.98, 95%CI=0.87-1.10, p=0.74) effect models (Table 2; Figure 2). Mutant heterozygous genotype (AG vs.GG; co-dominant model) meta-analysis did not show any significant association with BPD using fixed (OR_AG vs. GG_= 0.1.02, 95%CI= 0.95-1.07, p= 0.57) and random (OR_AG vs. GG_= 1.02, 95%CI= 0.93-1.13, p= 0.56) effect models (Table 2). Combined mutant genotypes (AA+AG vs. GG; dominant model) meta-analysis showed negative association with BPD using both fixed (OR_AA+AG vs. GG_= 1.0, 95%CI= 0.94-1.06, p= 0.89) and random (OR_AA+AG vs. GG_=1.0, 95%CI= 0.92-1.09, p= 0.86) effect models. Similarly the recessive genotypes model (AA vs. AG+GG) also did not show any association fixed (OR_AA vs. AG+GG_= 0.96, 95%CI= 0.89-1.05, p= 0.47) and random (OR_AA vs. AG+GG_= 0.96, 95%CI= 0.86-1.06, p= 0.45) effect models (Table 2).

**Table 2.** Summary estimates for the odds ratio (OR) in various allele/genotype contrasts, the significance level (p value) of heterogeneity test (Q test), and the I^2^ metric: overall analysis

| Genetic Models | Fixed effect<br>OR (95% CI), p | Random effect<br>OR (95% CI), p | Heterogeneity p-value (Q<br>test) | I<sup>2</sup> (%) | p value<br>of<br>Egger's<br>test) |
| --- | --- | --- | --- | --- | --- |
| Allele Contrast (A vs. G) | 0.99(0.94-1.03),0.49 | 0.98(0.93-1.03),0.46 | 0.02 | 34.64 | 0.09 |
| Co-dominant (AG vs. GG) | 1.02(0.95-1.07),0.57 | 1.02(0.93-1.13),0.56 | <0.0001 | 54.68 | 0.49 |
| Homozygote (AA vs. GG) | 0.98(0.89-1.08),0.75 | 0.98(0.87-1.10),0.74 | 0.13 | 20.43 | 0.05 |
| Dominant (AA+AG vs. GG) | 1.00(0.94-1.06),0.89 | 1.00(0.92-1.09),0.86 | 0.0007 | 46.83 | 0.81 |
| Recessive ( AA vs. GG+AG) | 0.96(0.89-1.05),47 | 0.96(0.86-1.06),0.45 | 0.09 | 23.92 | 0.02 |

**Figure 2.**
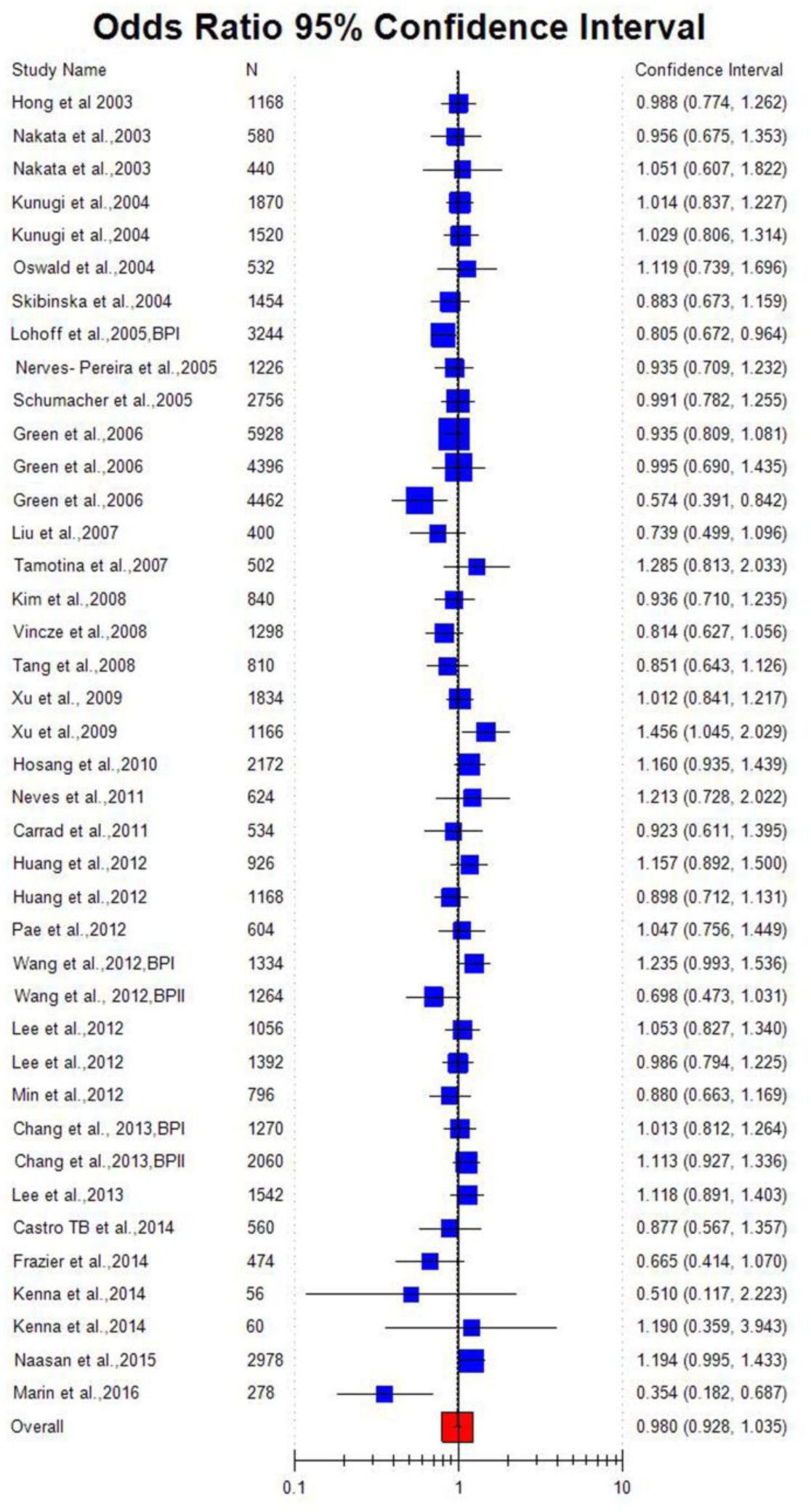
Random effect Forest plot of allele contrast model (A vs. G) of total 40 studies of BDNF Val66Met(G196A)polymorphism.

### Subgroup analysis

Out of 40 studies included in the present meta-analysis, 21 studies were carried out on Asian subjects, and 19 studies were carried out on Caucasian population. The subgroup analysis by ethnicity revealed that no significant association existed between BDNF Val66Met polymorphism and BPD in Asian population (OR_A vs.G_ = 1.02, 95%CI= 096-1.07., p=0.47; co-dominant model-OR_AG vs. GG_= 1.08, 95%CI= 0.99-1.18, p= 0.07; homozygote model- OR_AA vs. GG_ = 1.05, 95%CI=094-1.17., p=0.34; dominant model- OR_AA+AG vs. GG_= 1.07, 95%CI= 0.0.98-1.16, p= 0.09; recessive model-OR_AA vs. AG+GG_= 1.01, 95%CI= 0.92-1.11, p= 0.79) (Table 3, Figure 3). Similarly in Caucasian subgroup analysis, heterogeneity was low and except allele contrast model, significant association was not found between C677T polymorphism and BC risk (OR_A vs.G_ =0.93, 95%CI= 0.87-0.99, p=003.; co-dominant model-OR_AG vs. GG_= 0.96, 95%CI= 0.89-1.04, p= 0.34; homozygote model- OR_AA vs. GG_ = 0.80, 95%CI=0.65-0.98, p=0.03; dominant model- OR_AA+AG vs. GG_=0.94, 95%CI= 0.87-1.02, p= 0.14; recessive model-OR_AA vs. AG+GG_= 0.79, 95%CI= 0.65-0.95, p= 0.02) (Table 3,Figure 4).

**Table 3.** Summary estimates for the odds ratio (OR) in various allele/genotype contrasts, the significance level (p value) of heterogeneity test (Q test), and the I^2^ metric: subgroup analysis based on control source

| Genetic Models | Fixed effect<br>OR (95% CI), p | Random effect<br>OR (95% CI), p | Heterogeneity p-value (Q test) | I<sup>2</sup> (%) | p value of Egger's test) |
| --- | --- | --- | --- | --- | --- |
| **Asian studies ( studies)** |  |  |  |  |  |
| Allele Contrast (A vs. G) | 1.02(0.96-1.07),0.47 | 1.01(0.96-1.07),0.49 | 0.42 | 2.8 | 0.17 |
| Co-dominant (AG vs. GG) | 1.08(0.99-1.18),0.07 | 1.09(0.95-1.25),0.20 | 0.001 | 55.43 | 0.30 |
| Homozygote (AA vs. GG) | 1.05(0.94-1.17),0.34 | 1.05(0.94-1.17),0.34 | 0.56 | 0 | 0.60 |
| Dominant (AA+AG vs. GG) | 1.07(0.98-1.16),0.09 | 1.07(0.96-1.19),0.20 | 0.05 | 35.89 | 0.45 |
| Recessive ( AA vs. GG+AG) | 1.01(0.92-1.11),0.79 | 1.01(0.91-1.11),0.77 | 0.35 | 7.62 | 0.13 |
| **Caucasian studies ( studies)** |  |  |  |  |  |
| Allele Contrast (A vs. G) | 0.93(0.87-0.99),0.03 | 0.91(0.82-1.01),0.08 | 0.009 | 48.77 | 0.18 |
| Co-dominant (AG vs. GG) | 0.96(0.89-1.04),0.34 | 0.95(0.83-1.08),0.48 | 0.004 | 51.91 | 0.92 |
| Homozygote (AA vs. GG) | 0.80(0.65-0.98),0.03 | 0.77(0.57-1.04),0.09 | 0.07 | 33.89 | 0.12 |
| Dominant (AA+AG vs. GG) | 0.94(0.87-1.02),0.14 | 0.93(0.82-1.05),0.26 | 0.004 | 51.96 | 0.63 |
| Recessive ( AA vs. GG+AG) | 0.79(0.65-0.95),0.02 | 0.76(),0.05 | 0.09 | 31.66 | 0.07 |
| **BPD I studies ( studies)** |  |  |  |  |  |
| Allele Contrast (A vs. G) | 0.98(0.93-1.06),0.95 | 1.00(0.93-1.08),0.90 | 0.23 | 21.67 | 0.80 |
| Co-dominant (AG vs. GG) | 0.93(0.85-1.02),0.16 | 0.94(0.84-1.05),0.33 | 0.26 | 18.4 | 0.36 |
| Homozygote (AA vs. GG) | 1.06(0.91-1.23),0.43 | 1.06(0.91-1.23),0.43 | 0.77 | 0 | 0.76 |
| Dominant (AA+AG vs. GG) | 0.95(0.83-1.04),0.26 | 0.96(0.86-1.07),0.54 | 0.22 | 22.19 | 0.33 |
| Recessive ( AA vs. GG+AG) | 1.08(0.94-1.20),0.23 | 1.08(0.94-1.23),0.23 | 0.84 | 0 | 0.44 |
| **BPD II studies ( studies)** |  |  |  |  |  |
| Allele Contrast (A vs. G) | 1.10(1.01-1.13),0.04 | 1.08(0.93-1.14),0.04 | 0.29 | 16.58 | 0.85 |
| Co-dominant (AG vs. GG) | 1.31(1.06-1.60),0.03 | 1.3(1.1-1.7),0.02 | 0.14 | 30.23 | 0.52 |
| Homozygote (AA vs. GG) | 1.11(1.01-1.23),0.03 | 1.11(0.92-1.33),0.04 | 0.45 | 0 | 0.93 |
| Dominant (AA+AG vs. GG) | 1.21(1.06-1.37),0.003 | 1.21(1.01-1.46),0.03 | 0.07 | 42.67 | 0.27 |
| Recessive ( AA vs. GG+AG) | 0.96(0.83-1.12),0.06 | 0.97(0.80-1.19),0.08 | 0.17 | 28.93 | 0.73 |
| **BPD studies ( studies)** |  |  |  |  |  |
| Allele Contrast (A vs. G) | 0.95(0.89-1.02),0.24 | 0.93(0.85-1.02),0.15 | 0.06 | 37.12 | 0.0008 |
| Co-dominant (AG vs. GG) | 1.02(0.93-1.12),0.63 | 0.98(0.86-1.13),0.87 | 0.03 | 42.87 | 0.22 |
| Homozygote (AA vs. GG) | 0.82(0.68-0.98),0.03 | 0.76(0.62-1.01)0.07 | 0.07 | 35.97 | 0.06 |
| Dominant (AA+AG vs. GG) | 0.99(0.90-1.08),0.88 | 0.95(0.84-1.08),0.48 | 0.03 | 41.18 | 0.08 |
| Recessive ( AA vs. GG+AG) | 0.82(0.70-0.96),0.02 | 0.80(0.64-0.99),0.05 | 0.07 | 35.96 | 0.02 |

**Figure 3.**
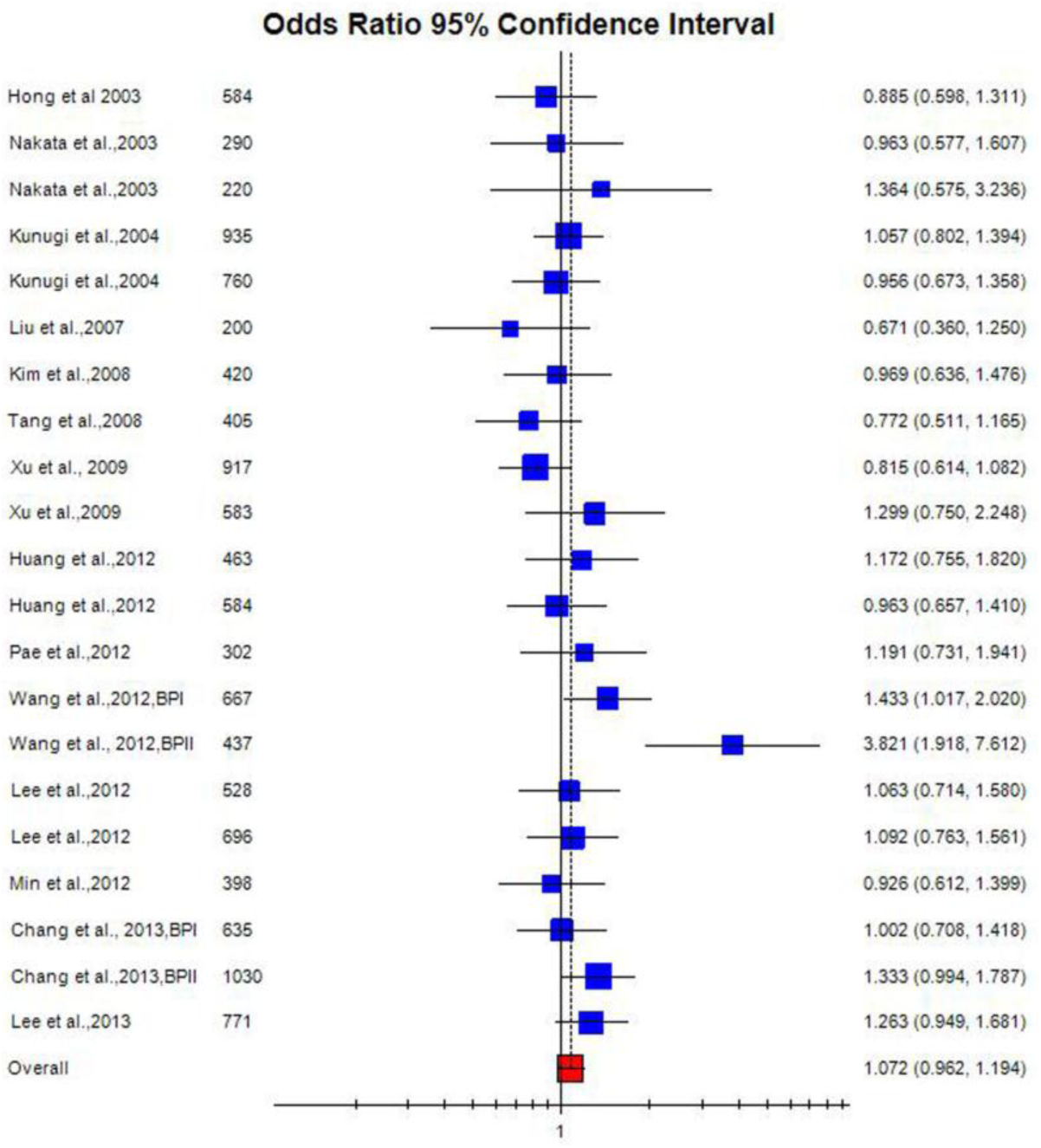
Random effect Forest plot of dominant model (AA+AG vs. GG) of 21 studies of Asian population of BDNF Val66Met(G196A)polymorphism.

**Figure 4.**
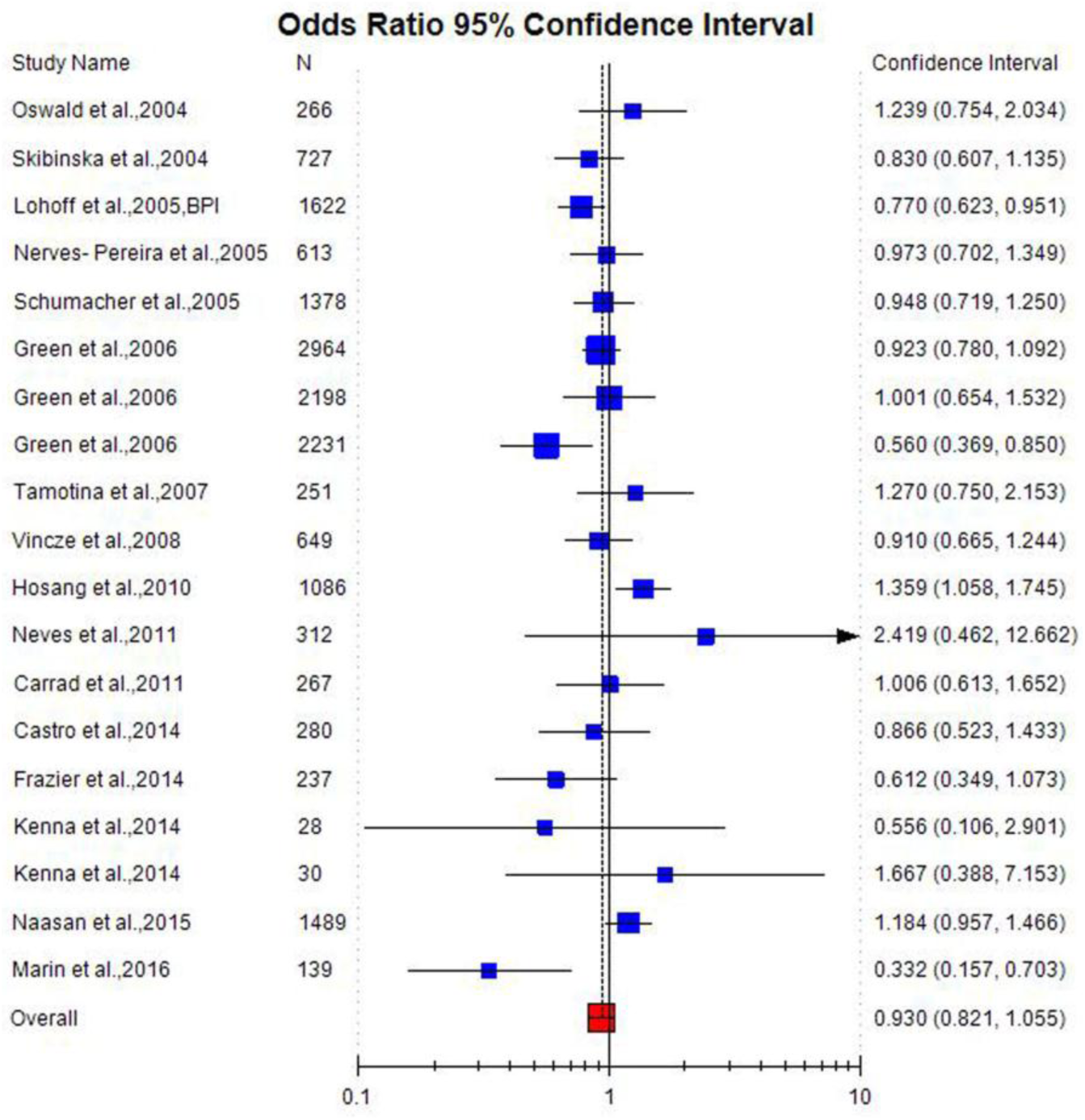
Random effect Forest plot of dominant model (AA+AG vs. GG) of 19 studies of Caucasian population of BDNF Val66Met(G196A)polymorphism.

Subgroup analysis based on subtypes of BPD like BPD I and BPDII was also performer. Out of 40 studied, in 12 studies cases were of BPD I and in 10 studies, authors selected BPD II cases for their analysis. In other studies, type of bipolar disorder was not mentioned, so we grouped them in one subgroup (BPD subgroup). Meta-analysis of BPD I studies did not show any association between Val66Met and BPD I (OR_A vs.G_ =0.98, 95%CI= 0.93-1.06, p=0.95; co-dominant model-OR_AG vs. GG_= 0.93, 95%CI= 0.85-1.02, p= 0.16; homozygote model- OR_AA vs. GG_ = 1.06, 95%CI=0.0.91, p=0.43; dominant model- OR_AA+AG vs. GG_=0.95, 95%CI= 0.83-1.04, p= 0.26; recessive model-OR_AA vs. AG+GG_=1.08, 95%CI= 0.0.94-1.20, p= 0.23) (Table 3). Meta-analysis of BPD II studies showed significant association between BDNF Val66Met polymorphism and BPDII except recessive model (OR_A vs.G_ = 1.10, 95%CI= 1.01, p=0.04; co-dominant model-OR_AG vs. GG_= 1.31, 95%CI= 1.06-1.60, p= 0.03; homozygote model- OR_AA vs. GG_ =1.11, 95%CI= 1.01-1.23, p=0.03; dominant model- OR_AA+AG vs. GG_= 1.21, 95%CI= 1.06-1.37, p= 0003; recessive model-OR_AA vs. AG+GG_=0.96, 95%CI= 0.83-1.12, p= 0.06) (Table 3).

Again subgroup BPD meta-analysis did not show any association between Val66Met polymorphism and BPD (OR_A vs.G_ =0.95, 95%CI= 0.89-1.02, p=0.24; co-dominant model-OR_AG vs. GG_= 1.02, 95%CI= 0.93-1.12, p= 0.63; homozygote model- OR_AA vs. GG_ = 0.82, 95%CI=0.68-0.98, p=0.03; dominant model- OR_AA+AG vs. GG_=0.99, 95%CI= 0.90-1.08, p= 0.88; recessive model-OR_AA vs. AG+GG_=82, 95%CI= 0.70-0.96, p= 0.02) (Table 3).

### Heterogeneity and Sensitivity analysis

Between studies heterogeneity existed moderate heterogeneity in allele contrast (P_heterogeneity_ =0.02, I^2^= 34.64), high heterogeneity in co-dominant (P_heterogeneity_ <0.0001, I^2^= 54.68), insignificant low heterogeneity in homozygote (P_heterogeneity_ =0.13, I^2^= 20.43), moderate heterogeneity in dominant (P_heterogeneity_ =0.0007, I^2^= 46.83) and low heterogeneity in recessive (P_heterogeneity_ =0.09, I^2^= 23.92) comparisons (Table 2).

Sensitivity analysis was performed by eliminating studies with small sample size (<100) and control population deviating from HWE. Control population of three studies was not in HWE (Nerves-Pereira et al.,2011; Wang et al.,2012; Lee et al.,2013) and heterogeneity was not decreased after exclusion of these studies (I^2^= 34.4%; p=0.02). Sample size of six studies was less than 100 (Nakata et al.,2003; Green et al.,2006; Xu et al.,2009;Wang et al.,2012; Kenna et al.,2014; Marin et al.,2016) and after exclusion of these studies heterogeneity was decreased (I^2^= 34.5%; p= 0.027).

### Publication bias

Funnel plots and Egger’s test were performed to estimate the risk of publication bias. Except recessive model, publication bias was absent (A vs. G: P Egger’s test= 0.09; AG vs. GG: P Egger’s test= 0.49; AA vs. GG: P Egger’s test P= 0.05; Dominant model AA+AG vs. GG: P _Egger’s test_= 0.81; Recessive model AA vs. AG+GG: P _Egger’s test_= 0.02).

## Discussion

Forty case–control studies with 10,085 bipolar disorder cases and 18,702 controls were included in the present meta-analysis. The results of current meta-analysis indicated that the Met (A) allele of BDNF was not associated with risk of BPD. Further, subgroup analyses based on ethnicity also did not show any association between BDNF Val66Met polymorphism and BPD in Asian as well as Caucasian populations. However, subgroup analysis based on BPD subtypes showed significant association with BPD II (OR_AA+AG vs. GG_= 1.21, 95%CI= 1.06-1.37, p= 0003) but not with BPD I. Although several evidences supported that BDNF factor is involved in BPD pathophysiology like- (i) reduced BDNF levels have been observed in patients with BPD during manic and depressive episodes and these levels have normalized with episode recovery (Cunha et al.,2006; Lin,2009), (ii) post-mortem studies have also demonstrated decreased hippocampal BDNF in patients with BPD (Knable et al.,2004; Dunham et al.,2009), (iii) use of antidepressants and mood stabilizers, e.g. lithium and valproate, to induce expression of neurotrophins (e.g., BDNF) and synaptic changes (Duman et al.,2000) and (iv) modulation of antidepressant-like affects by BDNF genotype and expression was also reported in human studies (Colle et al.,2015; Polyakova et al.,2015). Possible explanation for the lack of association between the Val66Met (rs6265) polymorphism and BPD in present meta-analysis may be due to (i) small sample size in included genetic association studies, (ii) different clinical criteria for selecting BPD patients and (iii) BDNF factor level decreases in subtype BPD II but not in BPD I.

Meta-analysis is an acceptable powerful statistical tool which is used effectively to combine data from several similar case control studies to obtain reliable results. During past two decades, numerous meta-analysis were published which evaluated genetic polymorphism as risk factor for different diseases and disorders- like-schizophrenia (Yadav et al., 2016; Rai et al.,2017), depression (Rai,2014), autism (Rai, 2016; Rai and Kumar,2018), Glucose 6-phosphate deficiency (Kumar et al.,2016), hyperuricemia (Rai, 2016), cleft lip and palate (Rai,2014,2017), male infertility (Rai and Kumar,2017), Down syndrome (Rai,2011; Rai et al., 2017; Rai and Kumar,2018), epilepsy (Rai and Kumar, 2018), Uterine Leiomyoma (Kumar and Rai, 2018), recurrent pregnancy loss (Rai, 2016), breast cancer (Rai,2014; Rai et al.,2017), digestive tract cancer (Yadav et al.,2018), colorectal cancer (Rai, 2015), esophageal cancer (Kumar and Rai,2018), ovary cancer (Rai,2016), and prostate cancer (Yadav et al., 2016) and endometrial cancer (Kumar and Rai,2018) etc.

Despite the clear strengths of present meta-analysis, including relatively large sample sizes and lack of publication bias, the interpretation of results should be done in light of few limitations like-(i) crude ORs without adjustment was used as association measure, adjusted analysis could not be done due to lack of sufficient raw data about related risk factors like substance abuse, alcohol intake etc., (ii) significant heterogeneity was observed in overall meta-analysis,(iii) single gene polymorphism was considered, and (iv) gene–environment interactions were not considered.

## Conclusions

We performed present meta-analysis to find out association between BDNF Val66Met polymorphism and bipolar disorder susceptibility. The results of present meta-analysis reported that BDNF Val66Met polymorphism is not risk factor for bipolar disorder (OR_A vs.G_ =0.99, 95%CI= 0.94-1.03, p=0.49). According to DSM IV bipolar disorder is a heterogeneous disease with many subtypes -BPD I, BPD II etc and results or present study also showed that BDNF Val66Met polymorphism may be involved in the pathogenesis of BPD by influencing the susceptibility of specific subtypes such as BP II (OR=1.21, 95% CI=1.06-1.37, p=0.003). Further in subgroup analysis showed no association between Val66Met polymorphism and bipolar disorder risk either from Asian or Caucasian population. In future, studies with larger sample sizes from different ethnic population are required to reach a definitive conclusion regarding this association. Also, it is necessary to take into consideration different inheritance patterns and the interaction of the BDNF gene with the environment.

## Data Availability

All data are reported in the paper.

## Abbreviations

BPD: Bipolar disorder;
BDNF: Brain derived neurotrophic factor;
Val66Met: Valine66Methionine;
OR: Odd ratio;
CI: Confidence interval

## Ethics Approval and consent to participants

In present study, we did not use any human sample, so there is no need of ethics approval and consent of participants.

## Consent for Publication

Yes

## Authors Contributions

VR and PK designed and wrote this study. VR performed the meta-analysis and FH and PK collected data.

